# Orbitofrontal-striatal structural alterations linked to negative symptoms at different stages of the schizophrenia spectrum

**DOI:** 10.1101/2020.04.07.20057166

**Authors:** Matthias Kirschner, André Schmidt, Benazir Hodzic-Santor, Achim Burrer, Andrei Manoliu, Yashar Zeighami, Yvonne Yau, Nooshin Abbasi, Anke Maatz, Benedikt Habermeyer, Aslan Abivardi, Mihai Avram, Felix Brandl, Christian Sorg, Philipp Homan, Anita Riecher-Rössler, Stefan Borgwardt, Erich Seifritz, Alain Dagher, Stefan Kaiser

**Author notes:** Corresponding Author: Matthias Kirschner, McConnell Brain Imaging Centre, Montréal Neurological Institute, McGill University, 3801 Rue University, Montréal QC, H3A 2B4 Canada. equal contribution (Alain Dagher, Stefan Kaiser).

## Abstract

Among the most debilitating manifestations of schizophrenia are negative symptoms such as anhedonia and apathy. Imaging studies have linked these symptoms to morphometric abnormalities in two brain regions implicated in reward and motivation: the orbitofrontal cortex (OFC) and ventral striatum. Negative symptoms generally are associated with reduced OFC thickness, while apathy specifically maps to reduced striatal volume. However, it remains unclear whether these tissue losses are a consequence of chronic illness and its treatment, or an underlying phenotypic trait. Here we use multicentre MRI data to investigate orbitofrontal-striatal abnormalities across the schizophrenia-spectrum from healthy populations with schizotypy, to unmedicated and medicated first-episode psychosis patients, and patients with chronic schizophrenia. Striatal volumes and OFC thickness were estimated from T1-weighted images acquired in all three diagnostic groups and controls from four sites (n=337). Results were first established in one test cohort (“Zurich sample”) and replicated in three independent samples. There was a correlation between apathy and striatal volume only in healthy individuals with schizotypy; however, medicated patients exhibited larger striatal volumes, which appears to be a consequence of antipsychotic medications. The association between reduced OFC thickness and negative symptoms generally also appeared to vary along the disease course, being significant only in first-episode psychosis patients. In schizotypy there was increased OFC relative to controls. Our findings suggest that negative symptoms associate with a temporal continuum of orbitofrontal-striatal abnormalities that may predate the occurrence of schizophrenia. Thicker OFC in schizotypy may represent either compensatory or pathological mechanisms prior to disease-onset.

## Introduction

Negative symptoms of schizophrenia are a multifaceted clinical construct occurring in all stages of the disorder.^1^ Consistent with a continuum model of psychosis, subclinical manifestations of negative symptoms can also be observed in the general population, especially in individuals with higher psychosis-proneness such as schizotypy.^2–4^ Negative symptoms comprise a diverse set of symptoms including avolition, anhedonia, asociality, alogia and blunted affect.^5,6^ In particular motivational and hedonic deficits (herein referred to as apathy) are associated with diminished quality of life, higher disease burden and in general poor long-term prognosis.^7–11^ Converging lines of research emphasize that functional and structural abnormalities of the cortico-striatal reward circuit are involved in the development of negative symptoms in schizophrenia.^12–14^ One of the most consistent findings from functional magnetic resonance imaging (fMRI) in schizophrenia is the association between negative symptoms and blunted ventral striatum (VS) activation.^15,16^ More specifically, apathy has been related to blunted striatal activation during: reward anticipation^17–22^, reward feedback processing^23^ and prediction error coding.^24,25^ These associations extend across a continuum from the general population to individuals in at-risk stages and early psychosis^26,27^, although negative^28,29^ and contradictory findings have been reported.^30^ In addition, prefrontal hypoactivation has been linked to negative symptoms in patients with schizophrenia (SZ).^20,23–25,31,32^

Consistent with this accumulating evidence of abnormal fronto-striatal circuit function, a picture of neuroanatomical changes related to negative symptoms in schizophrenia has started to emerge.^12,33,34^ Two recent studies found that reduced VS volume is associated with apathy,^35,36^ while a recent meta-analysis found consistent evidence for an association between reduced orbitofrontal cortical thickness and global negative symptoms.^37^ Together, these findings suggest that structural correlates of negative symptoms in schizophrenia map onto a similar circumscribed set of brain regions implicated in motivation and reward. However, it is unclear whether this also holds across non-clinical and early stages of the schizophrenia-spectrum. Only few studies have directly investigated the association between structural brain abnormalities and negative symptoms in other groups of the schizophrenia-spectrum such as non-clinical populations with psychotic-like experiences (e.g. schizotypy)^38,39^, individuals in at-risk stages^40^, first-episode psychosis patients (FEP)^41,42^ or untreated schizophrenia.^43^ In this regard, transdiagnostic approaches may help to shed light on shared or divergent structural signatures of negative symptoms across the entire disease spectrum. Specifically, including non-clinical populations with high schizotypal personality traits (SPT) and only minimally treated first-episode psychosis patients offers the unique opportunity to study brain-behaviour relationships in the absence of structural changes related to long-term antipsychotic treatment and chronic psychosis.^44–46^ Thus, transdiagnostic approaches may inform whether tissue losses are a consequence of chronic illness and its treatment, or an underlying phenotypic trait such as negative symptoms.

Here, we used multicentre MRI data from four research centers to investigate orbitofrontal-striatal structural correlates of apathy and global negative symptoms across a continuum of non-clinical, unmedicated individuals with SPT, unmedicated and medicated patients with FEP, and patients with SZ. Based on previous findings in SZ^35–37^, we hypothesized that reduced striatal volumes are correlated with apathy, while reduced OFC thickness is correlated with global negative symptom severity within each subgroup of the schizophrenia-spectrum. We first performed the analysis in a dataset from Zurich that includes all three subgroups (SPT,FEP,SZ). We then attempted to confirm significant findings in three additional independent datasets including one sample of individuals with SPT (Montreal dataset), one sample of patients with FEP (Basel dataset), and one sample of patients with SZ (Munich dataset). Finally, to identify patterns of orbitofrontal-striatal structural abnormalities related to different stages of the schizophrenia-spectrum, we combined all four datasets (n=337) and compared orbitofrontal-striatal structural measures between subgroups of the schizophrenia-spectrum and healthy controls (HC).

## Methods

### Participants

#### Study I (Zurich dataset)

The current sample of 129 individuals (28HC, 27SPT, 26FEP, 48SZ) was derived from previously published fMRI studies investigating the association between reward system dysfunction and negative symptoms.^17,19,30^ All SZ patients and FEP patients were recruited from outpatient and inpatient units at the Psychiatric Hospital of the University of Zurich. Healthy individuals were recruited using an online survey of the Schizotypal Personality Questionnaire (SPQ).^47^ 956 participants completed the questionnaire (mean=16.66, SD=11.34). Individuals with the highest SPQ total scores (>10%) were invited to participate in the MRI study.

#### Study II (Montreal dataset)

The confirmation sample for the schizotypy group was derived from a neuroimaging study by Soliman et al.^48^ investigating high positive SPT based on the Perceptual Aberration Scale (PerAb)^49^ and high negative SPT based on the Physical Anhedonia Scale (PhysAn)^50^. Individuals were recruited from undergraduate classes at McGill University and SPT groups were recruited by virtue of scoring >1.95 S.D. above the mean of their sex on either scale but not on both. The total sample comprises 40 healthy individuals (13 high positive SPT, 12 high negative SPT, 15 low SPT).^48^

#### Study III (Basel dataset)

The confirmation sample for the FEP group was derived from previous neuroimaging studies.^51–53^ The initial sample comprises in total 119 individuals (74FEP, 45HC). 14FEP had to be excluded from the correlation analysis due to missing negative symptoms scores.

#### Study IV (Munich dataset)

The confirmation sample for the SZ group was derived from a study by Avram and colleagues (n=49).^54^ Twenty-six chronic SZ patients (>= two psychotic episodes and currently in symptomatic remission of positive symptoms according to^55^) and 23 HC were included.

All four studies were approved by the local ethics committees, and all participants gave written informed consent after receiving a complete description of the study. For more details on inclusion and exclusion criteria of all datasets please see supplementary methods.

### Image acquisition and processing

Image acquisition parameters are described in the supplementary methods. We processed T1-weighted structural brain scans from all four datasets using FreeSurfer Version 6.0.^56^ Based on our a priori hypothesis, we followed a region-of-interest (ROI) approach and extracted striatal volumes for the left and right nucleus accumbens (NAcc), caudate and putamen^57^, as well as left and right medial orbitofrontal cortex (MOFC) and lateral orbitofrontal cortex (LOFC) thickness values based on the Desikan–Killiany atlas.^58^ In total, our ROI analysis consists of 10 ROIs (5 for each hemisphere). Quality control followed the standardized ENIGMA protocols for subcortical and cortical analysis (http://enigma.ini.usc.edu/protocols/imaging-protocols), which have been used in several previous studies (see supplementary methods).^37,44,59–61^

### Assessment of Negative Symptoms

#### Study I (Zurich dataset)

The Brief Negative Symptoms Scale (BNSS) was used to assess the severity of negative symptoms in all individuals of the schizophrenia-spectrum (SPT,FEP,SZ).^62–64^ The BNSS total score combined all items and was used to measure global severity of negative symptoms. The BNSS apathy dimension (avolition, asociality, anhedonia) and BNSS diminished expression (blunted affect, alogia) was calculated following the 2-factor solution proposed by Mucci and colleagues.^65^ In this context BNSS apathy reflects reduced motivation and engagement in goal-directed behaviour and diminished experience of pleasure, while BNSS diminished expression reflects decrease in facial and vocal expression, or body gestures, and reduced quantity of speech and spontaneous elaboration.^5,66^ Of note, recent studies have suggested that a more fine-grained differentiation of negative symptoms into five factors may provide a better fit to the psychometric data.^67–69^ Here, we used the consensual two-factor/domain model, for the following reasons. First, psychometric advantages of a five-factor model across all groups of the schizophrenia-spectrum remain to be determined.^67^ Second, our a priori hypothesis is based on studies that have successfully applied two-factor/domain models to identify specific brain correlates of apathy.^17,19,36,70^ Third, a more granular differentiation into avolition, asociality, and anhedonia would increase the risk of type I error.

#### Study II (Montreal dataset)

The dataset from Soliman and colleagues^48^ did not include a clinical-rating of negative symptoms but used the 61 items from the PhysAn scale^50^ to assess negative schizotypy. PhysAn total scores were used to measure self-reported deficits in motivation and pleasure.

#### Study III (Basel dataset)

Negative symptoms in FEP were measured with the 24-item version of the Scale for the Assessment of Negative Symptoms (SANS).^71^ The Global SANS (summary) score consisted of all four global items (7,12,16,21). Following the approach from Strauss and colleagues ^10^, the SANS Global avolition-apathy subscore (items:16,21) was defined to assess deficits in motivation and pleasure and the SANS Global affect-alogia subscore (items:7,12) was defined to assess diminished expression.

#### Study IV (Munich dataset)

The Positive and Negative Symptom Scale (PANSS) was used and global negative symptoms were defined according to the Marder negative symptom factor.^72^ To assess severity of apathy, we calculated the PANSS amotivation factor described previously.^11^

### Statistical analysis

#### Association with negative symptoms

For each subgroup (SPT,FEP,SZ) of the schizophrenia-spectrum, separate Spearman rank correlations were used to investigate an association of apathy and global/total negative symptoms with striatal volumes and OFC thickness (psych package 1.8.12^73^, R version 3.6.1^74^). Based on previous findings^35–37^, an association between apathy and reduced striatal volume and an association between global negative symptoms and reduced OFC thickness were hypothesized. Covariates were included following the statistical approach of recent studies from the ENIGMA consortium.^37,44,59,61^ For subcortical analysis the model included age, sex and intracranial volume (ICV) as covariates. For cortical analyses, the model included age and sex as covariates. In order to confirm significant findings from study I, the same correlation analyses were repeated for healthy individuals with SPT using the dataset from study II, for FEP patients using the dataset from study III and for patients with SZ using the dataset from study IV. Multiple comparison of the primary analysis in study I was adjusted for the number of total tests (n=30, 10 tests per each group) using false discovery rate p<0.05 (Benjamini– Hochberg procedure).^75^ Subsequent confirmation analyses in study II, study III and study IV were only performed in those ROIs in which significant association with either apathy and subcortical volumes or global negative symptoms and OFC thickness had been observed. Confirmation analyses are reported with an uncorrected threshold of p<0.05.

#### Group comparison

Group comparisons with data from all sites (n=337) were performed using multivariate analysis of covariance (MANCOVA) and univariate analysis of covariance (ANCOVA) for subsequent post-hoc analyses (jmv package^76^,R version 3.6.1^74^). Following recent reports^37,44,59,61^, two separate models were defined: (1) for striatal volumes including accumbens, caudate and putamen volume as dependent variable, group as fixed factor and age, sex, ICV, and acquisition site as covariates; (2) for cortical thickness including left and right MOFC, LOFC as dependent variable, group as fixed factor and age, sex, and site as covariates. Post-hoc pairwise comparisons were controlled for multiple comparison using the Bonferroni correction.

## Results

### Demographics and clinical data

Demographic and clinical data from all four datasets can be found in Table 1.

**Table 1.**
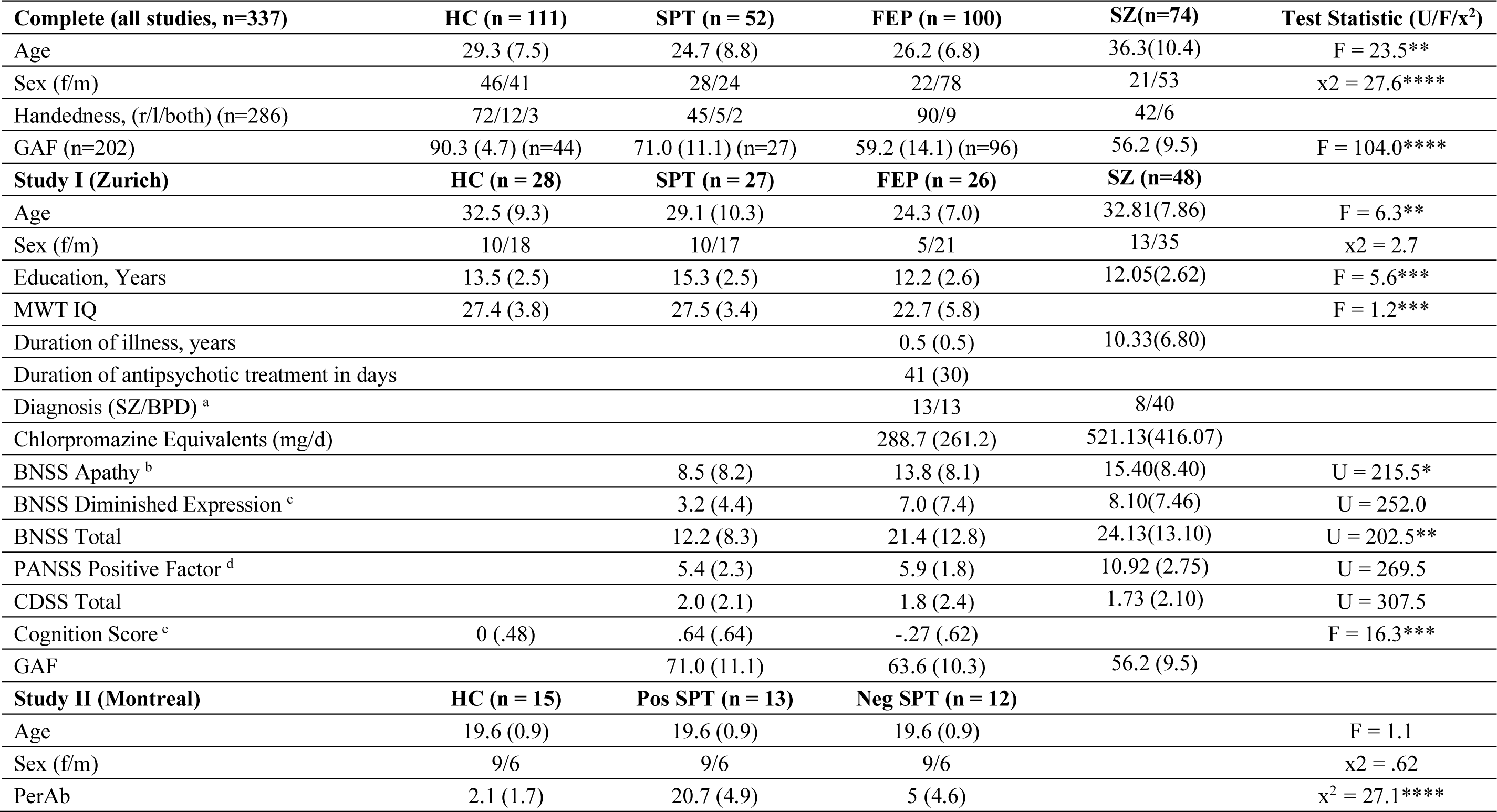
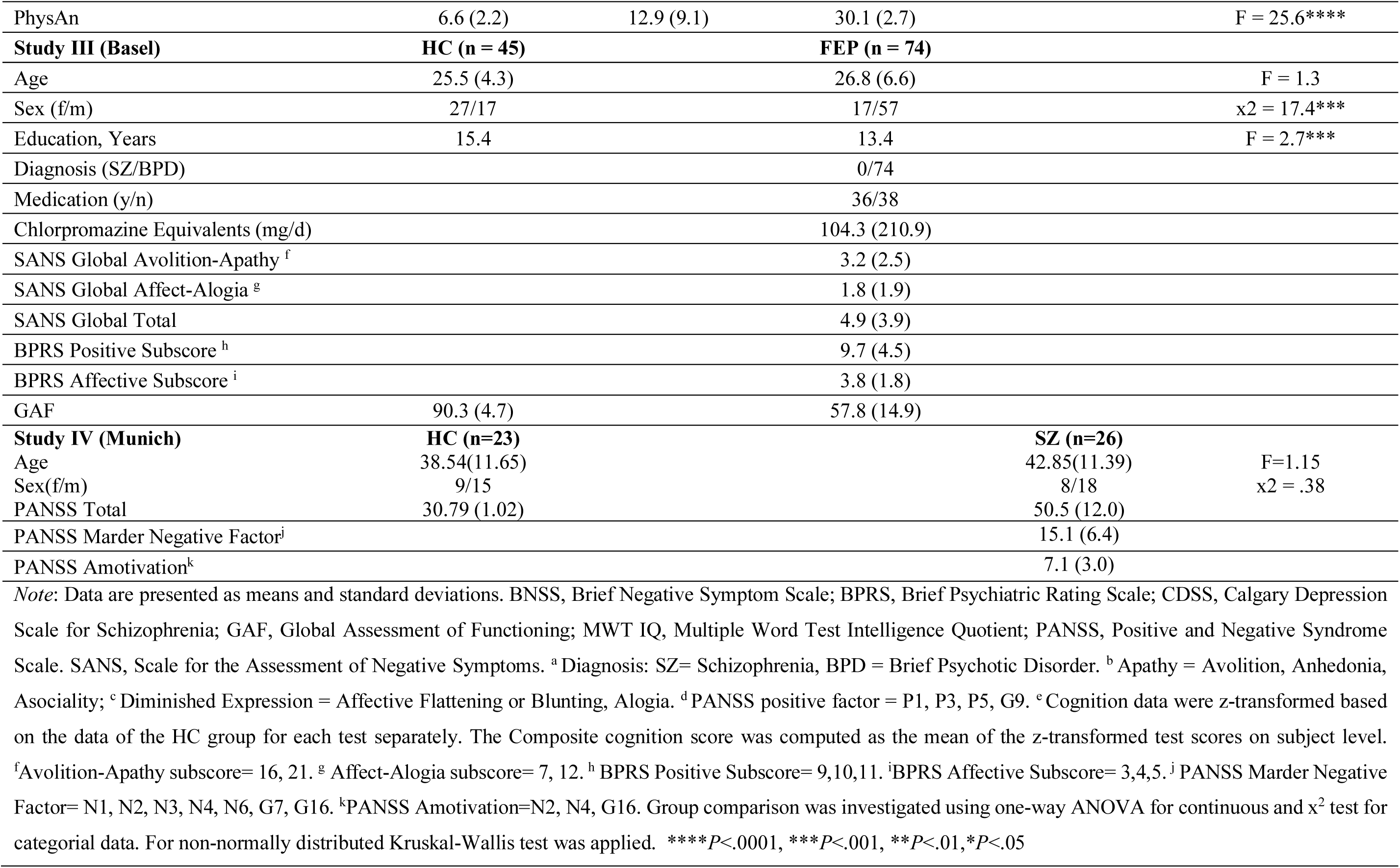
Demographic, Psychopathological and Clinical Data

| **Complete (all studies, n=337)** | **HC (n = 111)** | **SPT (n = 52)** | **FEP (n = 100)** | **SZ(n=74)** | **Test Statistic (U/F/x<sup>2</sup>)** |
| --- | --- | --- | --- | --- | --- |
| Age | 29.3 (7.5) | 24.7 (8.8) | 26.2 (6.8) | 36.3(10.4) | F = 23.5** |
| Sex (f/m) | 46/41 | 28/24 | 22/78 | 21/53 | x<sup>2</sup> = 27.6**** |
| Handedness, (r/l/both) (n=286) | 72/12/3 | 45/5/2 | 90/9 | 42/6 |  |
| GAF (n=202) | 90.3 (4.7) (n=44) | 71.0 (11.1) (n=27) | 59.2 (14.1) (n=96) | 56.2 (9.5) | F = 104.0**** |
| **Study I (Zurich)** | **HC (n = 28)** | **SPT (n = 27)** | **FEP (n = 26)** | **SZ (n=48)** |  |
| Age | 32.5 (9.3) | 29.1 (10.3) | 24.3 (7.0) | 32.81(7.86) | F = 6.3** |
| Sex (f/m) | 10/18 | 10/17 | 5/21 | 13/35 | x<sup>2</sup> = 2.7 |
| Education, Years | 13.5 (2.5) | 15.3 (2.5) | 12.2 (2.6) | 12.05(2.62) | F = 5.6*** |
| MWT IQ | 27.4 (3.8) | 27.5 (3.4) | 22.7 (5.8) |  | F = 1.2*** |
| Duration of illness, years |  |  | 0.5 (0.5) | 10.33(6.80) |  |
| Duration of antipsychotic treatment in days |  |  | 41 (30) |  |  |
| Diagnosis (SZ/BPD) <sup>a</sup> |  |  | 13/13 | 8/40 |  |
| Chlorpromazine Equivalents (mg/d) |  |  | 288.7 (261.2) | 521.13(416.07) |  |
| BNSS Apathy <sup>b</sup> |  | 8.5 (8.2) | 13.8 (8.1) | 15.40(8.40) | U = 215.5* |
| BNSS Diminished Expression <sup>c</sup> |  | 3.2 (4.4) | 7.0 (7.4) | 8.10(7.46) | U = 252.0 |
| BNSS Total |  | 12.2 (8.3) | 21.4 (12.8) | 24.13(13.10) | U = 202.5** |
| PANSS Positive Factor <sup>d</sup> |  | 5.4 (2.3) | 5.9 (1.8) | 10.92 (2.75) | U = 269.5 |
| CDSS Total |  | 2.0 (2.1) | 1.8 (2.4) | 1.73 (2.10) | U = 307.5 |
| Cognition Score <sup>e</sup> | 0 (.48) | .64 (.64) | -.27 (.62) |  | F = 16.3*** |
| GAF |  | 71.0 (11.1) | 63.6 (10.3) | 56.2 (9.5) |  |
| **Study II (Montreal)** | **HC (n = 15)** | **Pos SPT (n = 13)** | **Neg SPT (n = 12)** |  |  |
| Age | 19.6 (0.9) | 19.6 (0.9) | 19.6 (0.9) |  | F = 1.1 |
| Sex (f/m) | 9/6 | 9/6 | 9/6 |  | x<sup>2</sup> = .62 |
| PerAb | 2.1 (1.7) | 20.7 (4.9) | 5 (4.6) |  | x<sup>2</sup> = 27.1**** |

|  |  |  |  |  |
| --- | --- | --- | --- | --- |
| PhysAn | 6.6 (2.2) | 12.9 (9.1) | 30.1 (2.7) | F = 25.6**** |
| **Study III (Basel)** | **HC (n = 45)** |  | **FEP (n = 74)** |  |
| Age | 25.5 (4.3) |  | 26.8 (6.6) | F = 1.3 |
| Sex (f/m) | 27/17 | | 17/57 | $\chi^2 = 17.4***$ |
| Education, Years | 15.4 |  | 13.4 | F = 2.7*** |
| Diagnosis (SZ/BPD) |  |  | 0/74 |  |
| Medication (y/n) |  |  | 36/38 |  |
| Chlorpromazine Equivalents (mg/d) |  |  | 104.3 (210.9) |  |
| SANS Global Avolition-Apathy <sup>f</sup> |  |  | 3.2 (2.5) |  |
| SANS Global Affect-Alogia <sup>g</sup> |  |  | 1.8 (1.9) |  |
| SANS Global Total |  |  | 4.9 (3.9) |  |
| BPRS Positive Subscore <sup>h</sup> |  |  | 9.7 (4.5) |  |
| BPRS Affective Subscore <sup>i</sup> |  |  | 3.8 (1.8) |  |
| GAF | 90.3 (4.7) |  | 57.8 (14.9) |  |
| **Study IV (Munich)** | **HC (n=23)** |  | **SZ (n=26)** |  |
| Age | 38.54(11.65) |  | 42.85(11.39) | F=1.15 |
| Sex(f/m) | 9/15 | | 8/18 | $\chi^2 = .38$ |
| PANSS Total | 30.79 (1.02) |  | 50.5 (12.0) |  |
| PANSS Marder Negative Factor<sup>j</sup> |  |  | 15.1 (6.4) |  |
| PANSS Amotivation<sup>k</sup> |  |  | 7.1 (3.0) |  |

*Note:* Data are presented as means and standard deviations. BNSS, Brief Negative Symptom Scale; BPRS, Brief Psychiatric Rating Scale; CDSS, Calgary Depression Scale for Schizophrenia; GAF, Global Assessment of Functioning; MWT IQ, Multiple Word Test Intelligence Quotient; PANSS, Positive and Negative Syndrome Scale. SANS, Scale for the Assessment of Negative Symptoms. <sup>a</sup> Diagnosis: SZ= Schizophrenia, BPD = Brief Psychotic Disorder. <sup>b</sup> Apathy = Avolition, Anhedonia, Asociality; <sup>c</sup> Diminished Expression = Affective Flattening or Blunting, Alogia. <sup>d</sup> PANSS positive factor = P1, P3, P5, G9. <sup>e</sup> Cognition data were z-transformed based on the data of the HC group for each test separately. The Composite cognition score was computed as the mean of the z-transformed test scores on subject level. <sup>f</sup>Avolition-Apathy subscore= 16, 21. <sup>g</sup> Affect-Alogia subscore= 7, 12. <sup>h</sup> BPRS Positive Subscore= 9,10,11. <sup>i</sup>BPRS Affective Subscore= 3,4,5. <sup>j</sup> PANSS Marder Negative Factor= N1, N2, N3, N4, N6, G7, G16. <sup>k</sup>PANSS Amotivation=N2, N4, G16. Group comparison was investigated using one-way ANOVA for continuous and $\chi^2$ test for categorical data. For non-normally distributed Kruskal-Wallis test was applied. \*\*\*\* $P < .0001$ , \*\*\* $P < .001$ , \*\* $P < .01$ , \* $P < .05$

### Association between apathy and reduced striatal volume

#### Individuals with SPT

In healthy individuals with SPT (n=27), BNSS apathy scores were negatively associated with volume of the right NAcc (rs=-.54, FDR-adjusted p=.02), and bilateral putamen (right: rs=-.58, FDR-adjusted p=.01: left: rs=-.63, FDR-adjusted p=.009) accounting for age, sex and intracranial volume (Fig. 1, Table 2A). Using Steiger’s test^77^, we could show that the associations between BNSS apathy with right NAcc and right putamen volume were significantly stronger than the association between BNSS apathy and the right caudate volume (Table S1). These findings indicate regional specificity of the relationship with apathy with respect to the right NAcc and right putamen volume.

**Table 2A.**
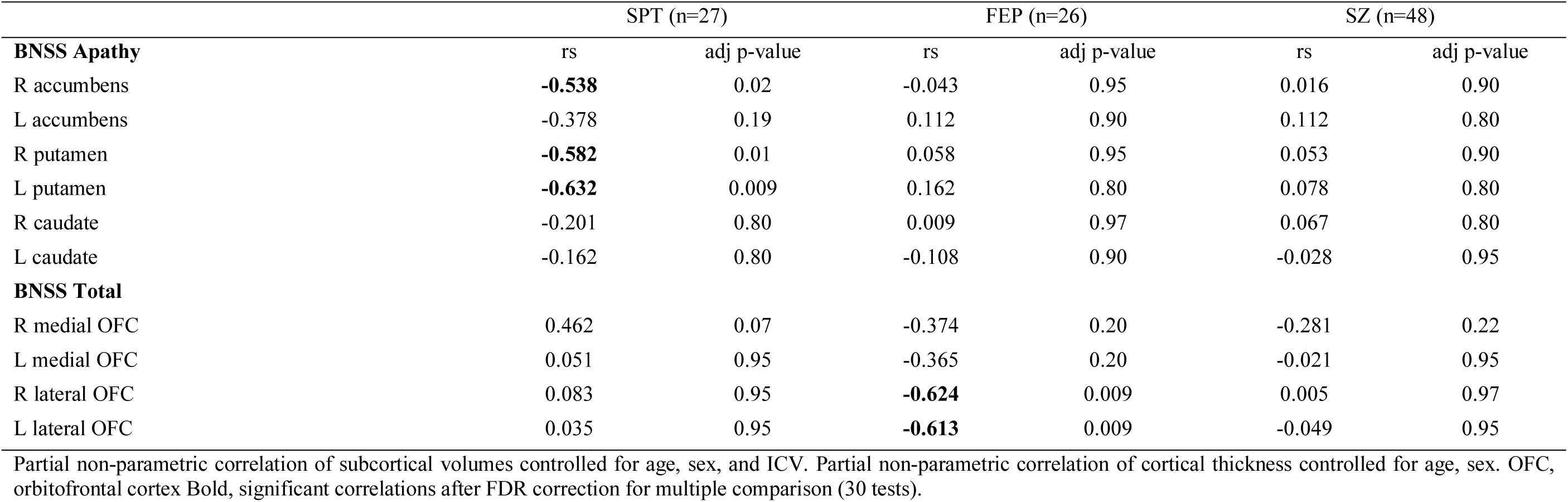
Spearman Correlation between Apathy and striatal ROIs and global negative symptoms and cortical ROIs

**Table 2B.**
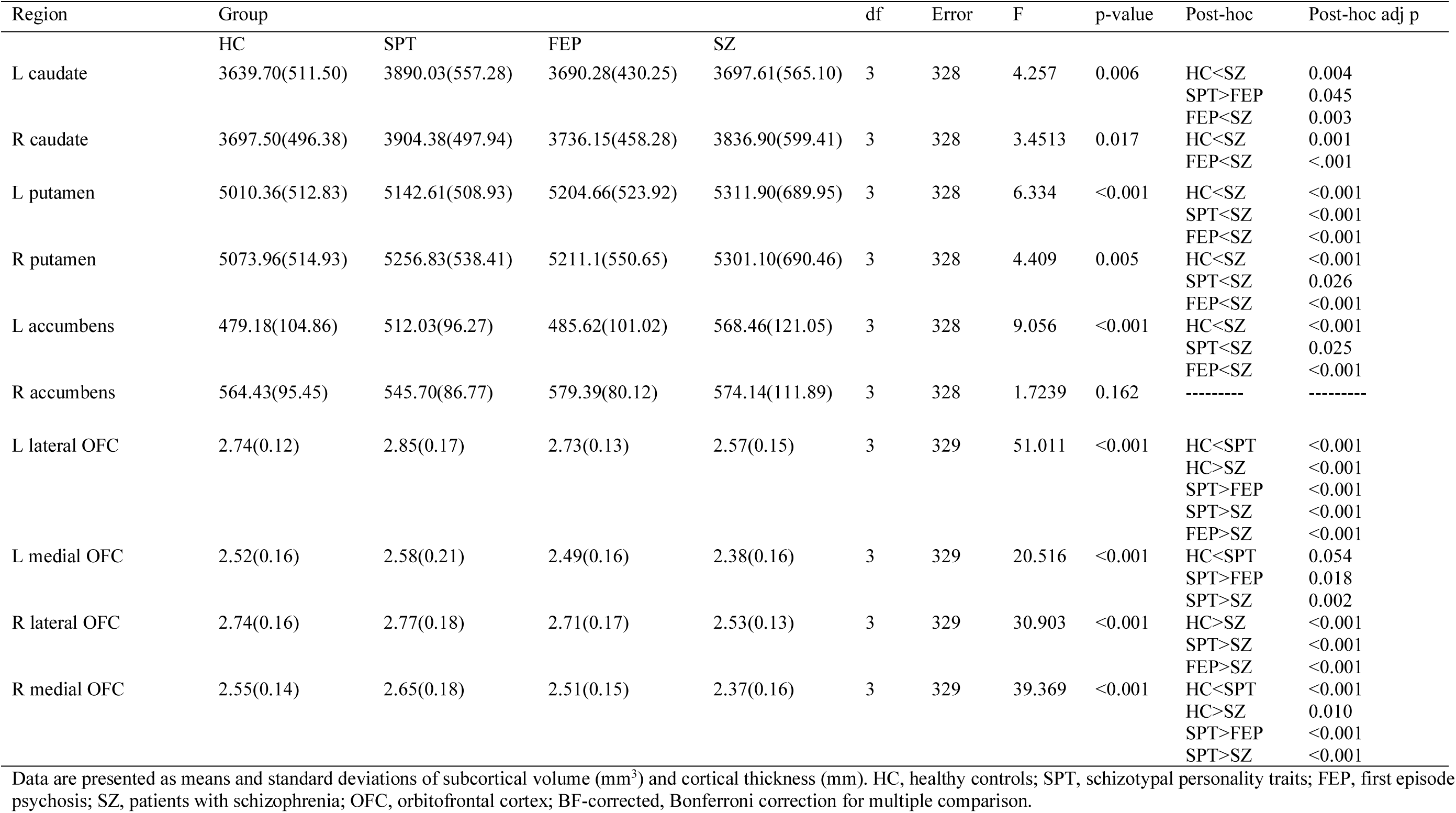
Group differences of subcortical volumes and OFC thickness

**Figure 1.**
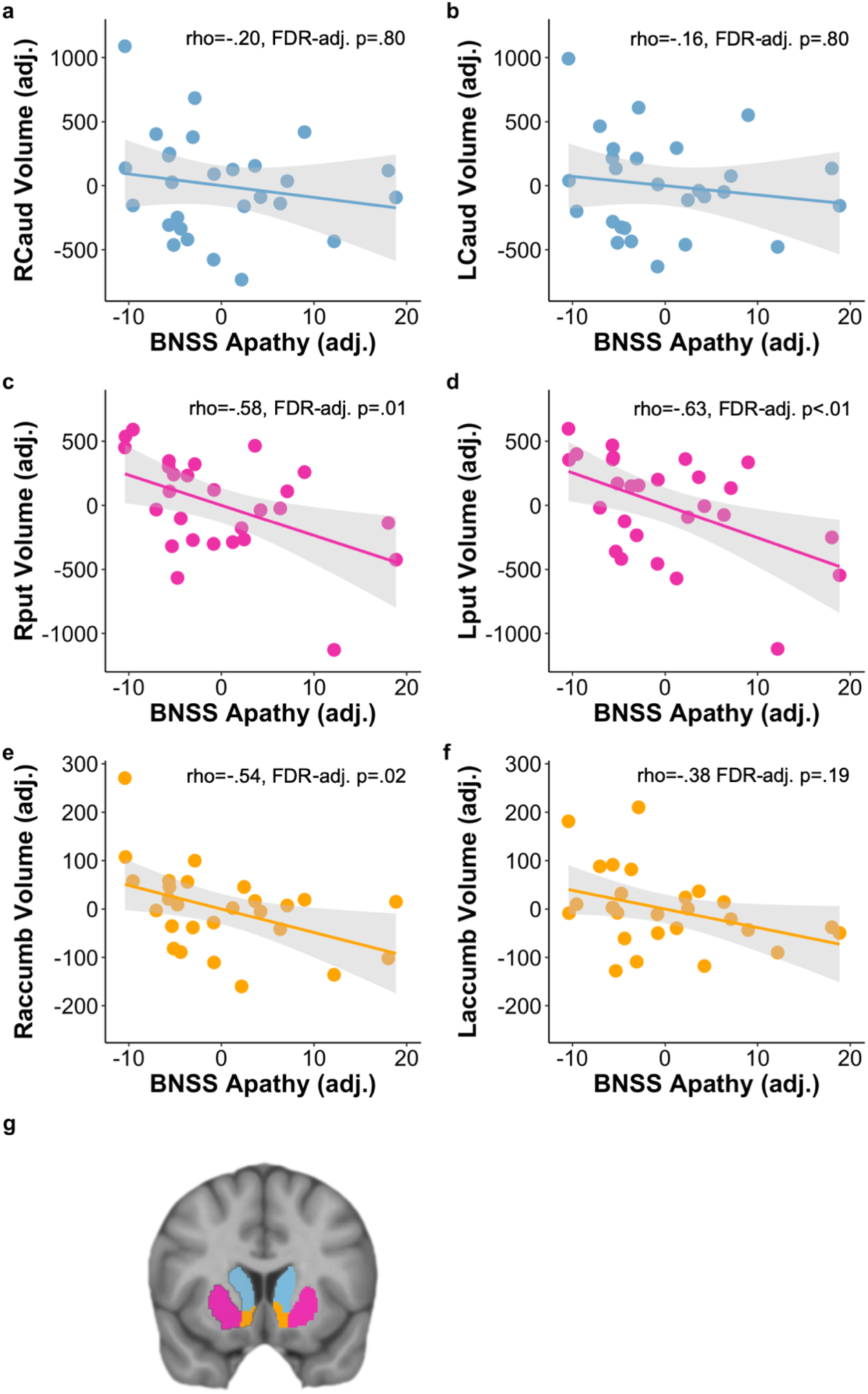
Association between striatal volumes and subclinical apathy in schizotypy. Zurich SPT Sample (n=27). For visualisation purpose, partial correlations between subcortical volume and BNSS Apathy are shown as Pearson correlation coefficients and two-tailed significance tests using R. Regressions were adjusted for age, sex, intracranial volume. Error shadings correspond to standard errors. adj., adjusted. Significant associations are observed in (c) right putamen (rs=-.58, FDR-adjusted p=.01), (d) left putamen (rs=-.63, FDR-adjusted p=.009) and (e) right NAcc (rs=-.54, FDR-adjusted p=.02). In contrast, correlation analysis between BNSS apathy and (a) right caudate volume (rs=-.20, FDR- adjusted p=.80), (b) left caudate volume (rs=-.16, FDR-adjusted p=.80), and (f) left NAcc (rs=-.38, FDR-adjusted p=.19) were not significant. (g) visualization of the six striatal volumes, skyblue=caudate, pink=putamen, orange= nucleus accumbens.

No association of subcortical volumes with BNSS diminished expression or potential confounding variables such as subclinical depressive symptoms, subclinical positive symptoms and cognition were observed (Table S1). Using Steiger’s test ^77^, we found that the associations between BNSS apathy and right NAcc as well as bilateral putamen were significantly stronger than the association between BNSS diminished expression and the respective striatal volumes (supplementary results). In other words, reduced striatal volumes are specifically associated with apathy and not related to other negative symptoms.

In a next step, we aimed to confirm these findings in an independent sample (Montreal dataset) with varying levels of schizotypy. We found a significant negative association of PhysAn with right and left NAcc and right putamen in the subgroup with positive schizotypy (n=13) and moderate severity of PhysAn (Raccumb: rs=-.84, p=.002; Laccumb: rs=-.68, p=.03; Rput: rs=- .78, p=.008). Pointing in the same direction, individuals with high negative schizotypy (n=12) showed a non-significant correlation between PhysAn right accumbens volume and right putamen volume (Raccumb: rs=-.495, p=.175, Rput: rs=-.318, p=.4) (Table S1). In addition, the subgroup with low positive and negative schizotypy (n=15) showed a significant negative correlation between PhysAn and right and left NAcc volumes (Raccumb: rs=-.51, p=.088; Laccumb: rs=-.62, p=.03) (Table S1, Fig S2).

### Patients with FEP and SZ

In FEP, BNSS apathy scores were not significantly associated with the volumes of striatal subregions (Table 2A). Similarly, BNSS apathy did not significantly correlate with striatal volumes in patients with SZ (Table 2A).

Taken together, reduced striatal volume was associated with different measures of motivation and pleasure (BNSS apathy, PhysAn) in two independent datasets of unmedicated healthy individuals with SPT. This relationship was not detectable in FEP and SZ.

### Association between global negative symptoms and OFC thickness

#### Individuals with SPT

In healthy individuals with SPT, global negative symptoms (BNSS Total) were not correlated with reduced OFC thickness. In contrast, greater right MOFC thickness was positively correlated with BNSS total scores (rs=0.462, p=0.02), but this effect did not remain significant after correction for multiple comparisons (Table 2A).

#### Patients with FEP and SZ

In patients with FEP, global negative symptoms (BNSS Total) were negatively associated with cortical thickness of right and left LOFC (right: rs=-.624, FDR-adjusted p=.02, left: rs=-.613, FDR-adjusted p=.02, Fig. 2, Table 2A). Pointing in the same direction, thickness of right and left MOFC showed trend-level negative correlations with BNSS total scores (Table 2A). Regarding potential confounding variables, OFC thickness was not associated with current medication dose (chlorpromazine equivalents), depressive symptoms, positive symptoms, and cognition (Table S4). Additionally, association between LOFC and global negative symptoms did not change when adding current medication dose as covariate in the model (right: rs=-.619, p=.002; left: rs=-.608, p=.002).

**Figure 2.**
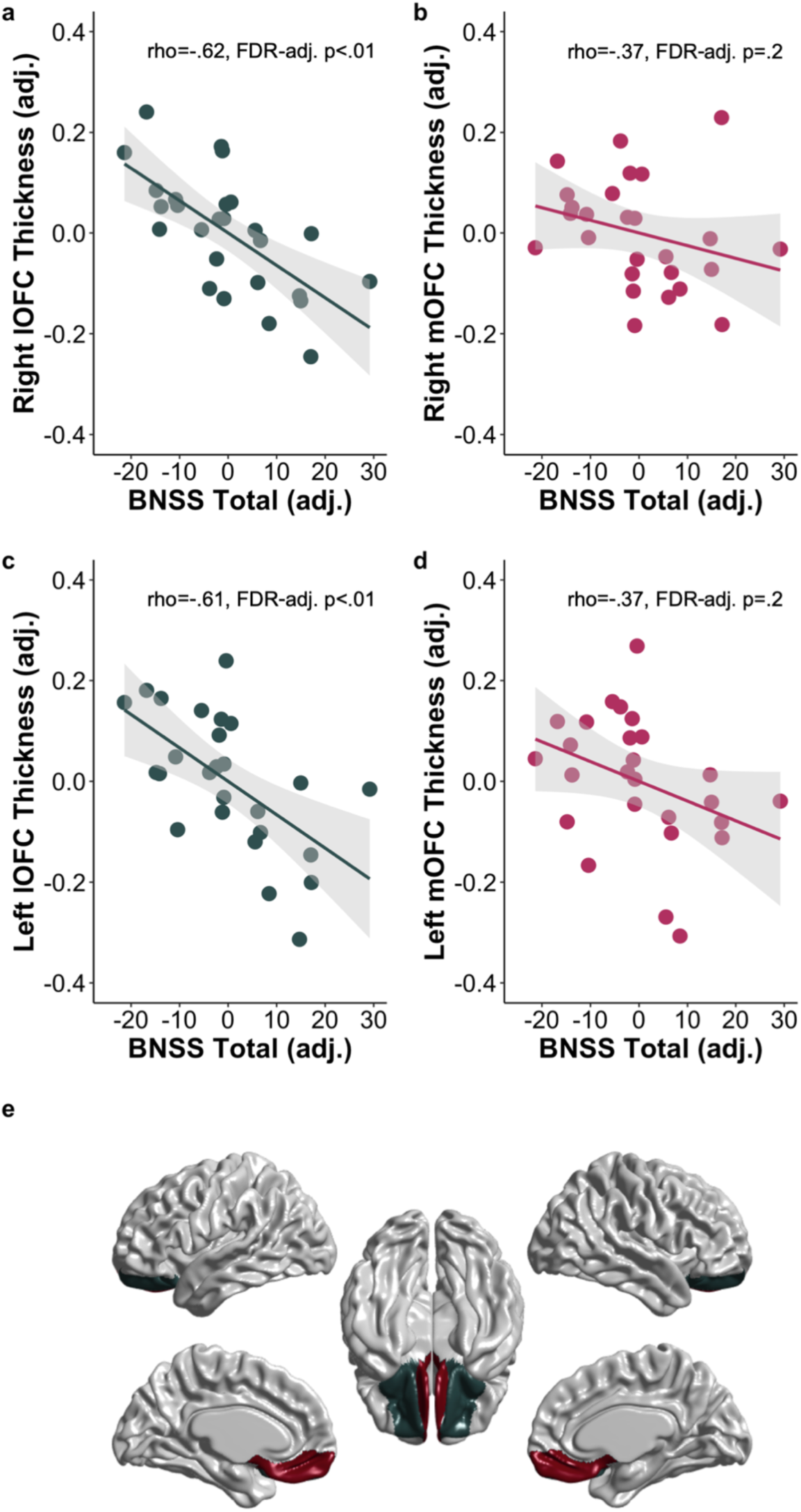
Association between cortical thickness and global negative symptoms in first-episode psychosis. FEP Zurich Sample (n=26). For visualisation purpose, partial correlations between OFC thickness and BNSS Total scores are shown as Pearson correlation coefficients and two-tailed significance tests using R. Regressions were adjusted for age, sex, intracranial volume. Error shadings correspond to standard errors. adj., adjusted. Significant associations are observed in (a) right LOFC (rs=-.624, FDR-adjusted p=.02) and (c) left OFC (rs=-.613, FDR-adjusted p=.02). Non-significant negative association are observed in (b) right MOFC (rs=-.624, FDR-adjusted p=.02) and (d) left MOFC (rs=-.624, FDR-adjusted p=.02). (e) visualization of OFC parcellations in surface space, green= lateral OFC, red= medial OFC.

In a next step, we aimed to confirm these findings in an independent FEP sample (n=60, Basel). We found a significant negative association of global negative symptoms (SANS global score) with left LOFC thickness (right: rs=-.13, p=.329, left: rs=-.319, p=.015). We addressed the relationship between OFC thickness and potential confounding variables and observed a significant correlation between depressive symptoms (BPRS depressive subscore) and left LOFC thickness (rs=-.27, p=.02) as well as depressive symptoms and global negative symptoms (rs=.51, p<.0001). Furthermore, when including depressive symptoms as additional covariate in the model the association between global negative symptoms and left LOFC was no longer significant (rs=-.169, p=.21). No association was observed with medication (chlorpromazine equivalents) or positive symptoms (Table S4) and the relationship between left LOFC and global negative symptoms did not change when adding current medication dose as covariate in the model (left: rs=-.338, p=.001).

Patients with SZ showed a non-significant negative correlation between right MOFC thickness and BNSS total scores (Table 2A).

In Sum, reduced LOFC thickness was associated with total negative symptoms in two independent FEP datasets. Conversely in SPT, MOFC thickness was positively correlated with subclinical negative symptoms.

### Group differences in striatal volume

We examined group differences of striatal volume and OFC thickness within the schizophrenia-spectrum across all studies (n=337). Using MANCOVA, we found a significant effect of group for striatal volumes (F(18,975)=6.66, p<.0001, Pillais Trace=0.329). Univariate analyses of covariance revealed significant group differences in bilateral caudate, putamen, and left accumbens (Fig.3 Table 2B). Post-hoc pairwise comparisons showed a general pattern of greater caudate, putamen and accumbens volumes in patients with SZ compared to all other groups (Fig.3, Table 2B). Thus, patients with the longest duration of illness and long-term antipsychotic treatment displayed significant enlargement of striatal volumes.

**Figure 3.**
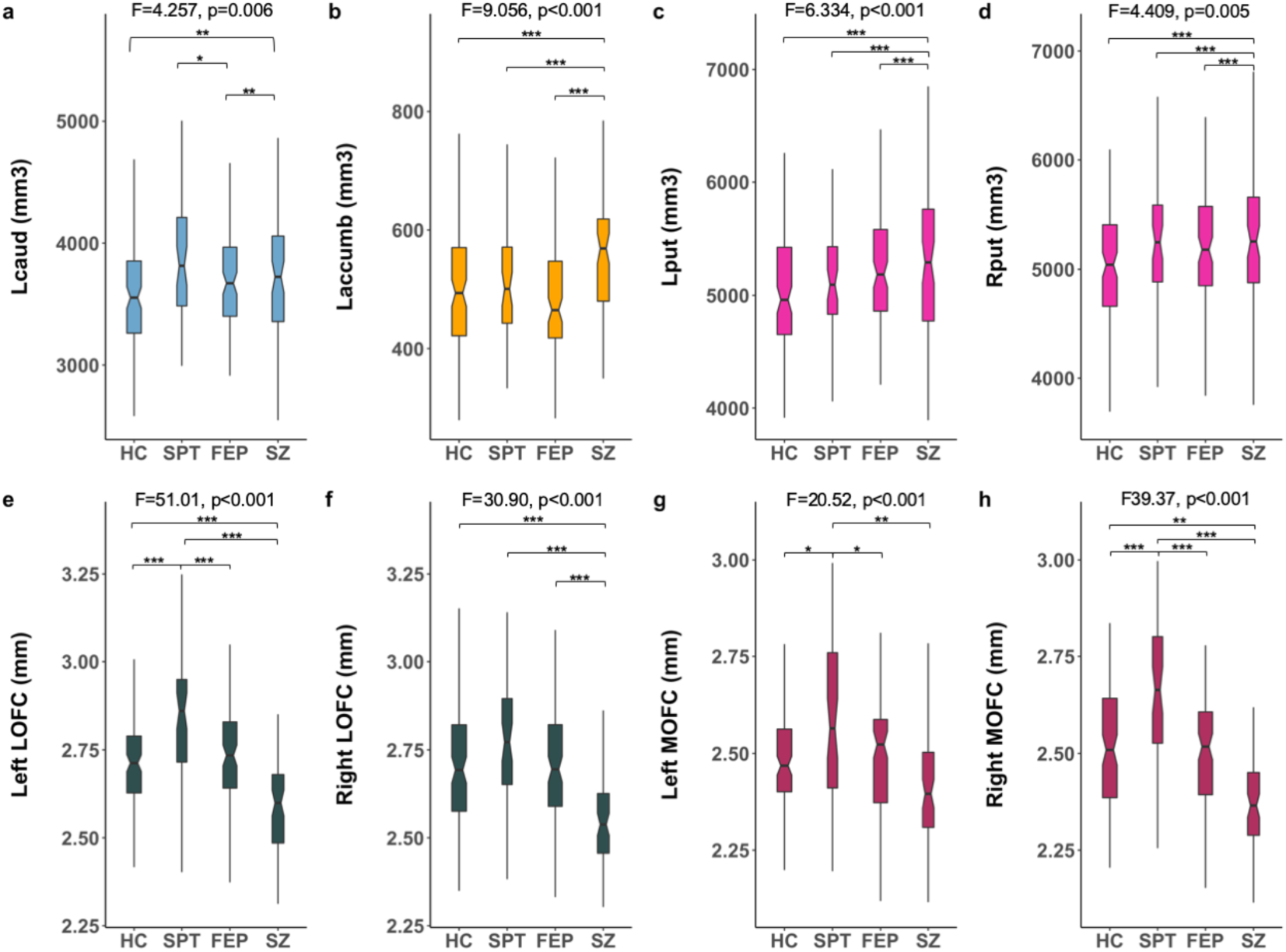
Group comparison of striatal volumes and OFC thickness across all sites (n=337) (a) Significant larger left caudate in SZ compared to HC and FEP (SZ>HC, p=.004, SZ>FEP, p=.003). (b) Significant larger left accumbens and (c,d) bilateral putamen volume in SZ compared to all other groups (SZ>HC, SZ>FEP, all ps<.001, SZ>SPT all ps<.03). (e) Significant greater left LOFC thickness in SPT compared to all other groups (all ps<.001) and greater left LOFC thickness in HC and FEP compared to SZ (all ps<.001). (f) Greater right LOFC thickness in HC, SPT, and FEP compared to SZ (HC>SZ, SPT>SZ, FEP>SZ, allps<.001). (g,h) Significant greater bilateral MOFC thickness in SPT compared to all other groups (all ps<.02) and greater right MOFC in HC compared to SZ (p=.01). All post-hoc group tests Bonferroni corrected for multiple comparison (p<.05). *<.05, **<.01, ***<.001.

To further explore effects of antipsychotic treatment on striatal volume in early disease stages, we divided the patients with FEP in one group of unmedicated FEP patients (FEP-NON) and one group of medicated FEP patients (FEP-MED) and performed an explorative post-hoc group comparison. FEP-MED (n=61) showed significant greater putamen volume (left: p=.007, right: p=.007) and left NAcc volume (Laccumb: p<.001, Raccumb: p=.067) compared to FEP-NON (n=39) (Table S3).

Taken together, medicated patients with SZ showed the greatest enlargement in striatal volumes compared to all other groups. Additionally, medicated FEP displayed greater putamen and left NAcc volume compared to unmedicated FEP.

### Group differences in OFC thickness

Regarding OFC thickness, MANCOVA revealed a significant effect of group for OFC thickness (F(12,984)=12.599, p<.001, Pillais Trace=.399). Univariate analyses of covariance revealed significant group differences in bilateral LOFC and MOFC (Fig.3, Table 2B). Post-hoc pairwise comparisons showed a general pattern of significantly thicker bilateral LOFC and MOFC in SPT compared to all other groups including HC. In addition, HC and FEP showed thicker OFC compared to SZ (Fig.3, Table 2B).

To further explore whether OFC thickness differences within the schizophrenia-spectrum might be related to onset of medication, we divided the FEP group in one unmedicated FEP patients’ group (FEP-NON) and one medicated FEP patients (FEP-MED) group and performed an explorative group comparison. FEP-NON showed significantly thicker MOFC (left: p=.002, right: p=.005) and LOFC (left: p=.001, right: p=.005) compared to FEP-MED (Table S4). Taken together, individuals with SPT displayed a pattern of thicker MOFC and left LOFC compared to all other groups (HC, FEP, SZ). In addition, FEP-NON showed thicker MOFC and LOFC compared to FEP-MED.

## Discussion

### Summary

This study revealed distinct orbitofrontal and striatal correlates of apathy and global negative symptoms across subgroups of the schizophrenia-spectrum. Reduced right NAcc and bilateral putamen volume correlated with apathy in unmedicated individuals with varying levels of SPT. This association was not observed in patient with FEP and SZ. In contrast, medicated patients (FEP,SZ) showed greater striatal volumes compared to unmedicated patients, SPT and HC. The relationship between OFC thickness and global negative symptoms revealed a different pattern. In patients with FEP, reduced LOFC thickness correlated with global negative symptoms. This association was not observed in unmedicated individuals with SPT, who showed an overall thicker OFC compared to all other groups including HC. In addition, medicated patients with FEP and SZ showed reduced OFC thickness compared to unmedicated FEP patients, unmedicated individuals with SPT and HC.

### Striatal volume

To our knowledge this is the first study demonstrating a relationship between severity of apathy and reduced striatal volume in unmedicated populations of the schizophrenia-spectrum. This association was even detectable in a small sample of healthy individuals with low schizotypal personality traits (Montreal dataset). An association of reduced ventral striatal volume with apathy has been previously reported for patients with SZ.^35,36^ Together with these previous studies, our results suggest that reduced striatal volumes are a specific neuroanatomical correlate of the apathy dimension but not the diminished expression dimension. Previous negative results, derived from analyses using global negative symptom scores^59,78^, could be explained by lack of differentiation into distinct negative symptom dimensions. With respect to other subgroups of the schizophrenia-spectrum little data are available yet. In contrast to the lack of structural MRI work, fMRI studies provide converging evidence for blunted striatal reward signal as dimensional and transdiagnostic correlate of motivational and hedonic deficits.^17,18,26,27,79^ Our findings in unmedicated individuals with subclinical motivational and hedonic deficits extend this work, providing first evidence for striatal structural changes in the pathophysiology of apathy prior to clinical manifestation.

Contrary to the existing literature on SZ^35,36^ and our observation in unmedicated individuals with SPT, apathy was not related to reduced striatal volume in medicated patients with schizophrenia-spectrum disorders (FEP,SZ). Of note, medicated SZ and FEP patients showed overall larger striatal volumes compared to healthy individuals and non-medicated patients. Accumulating evidence from human and animal studies show that long-term antipsychotic medication leads to subcortical grey matter enlargement.^80^ Are volumetric changes due to antipsychotic treatment or other disease-related processes responsible for our negative results? Within the schizophrenia-spectrum and compared to HC, medicated patients showed larger caudate, putamen and left accumbens volume. Observations of larger caudate and putamen volume (but not accumbens volume) in patients with SZ are consistent with two recent meta-analyses.^59,81^ Our results are also consistent with previous reports showing positive associations between larger striatal volumes and current medication dose as well as illness duration.^59,82,83^ In addition, longitudinal studies demonstrated that caudate and putamen volume increase after initiation of antipsychotic treatment in non-medicated or minimally treated patients.^84–87^ Our findings in FEP point in the same direction showing larger striatal volumes in medicated compared to unmedicated FEP patients. Although cross-sectional data do not allow causal inference, it is possible that in medicated patients associations between reduced striatal volumes and apathy are obscured by antipsychotic-induced volume changes.

### Orbitofrontal grey matter thickness

The present study revealed a dimensional association between global negative symptoms and reduced lateral OFC thickness in patients with FEP. The association between left lateral OFC thickness and global negative symptoms was replicated in an independent dataset of patients with FEP. In this regard, a recent meta-analysis provided evidence for an association between reduced medial and lateral OFC thickness and global negative symptoms in chronic SZ.^37^ Our data contribute and extend this finding demonstrating a dimensional relationship between OFC thickness and negative symptoms in FEP patients and a trend level effect in SZ patients. Importantly, this association was apparent in a mixed FEP sample (brief psychotic disorder and first-episode SZ, Zurich Dataset) and also in a minimally treated brief psychotic disorder sample (Basel Dataset). These results are consistent with two previous reports in smaller samples comparing patients with persistent (n<20) and non-persistent negative symptoms (n<55).^41,42^ Together with these studies, our data suggest that the association between orbitofrontal structural abnormalities and negative symptoms can occur early in the disease process prior to the diagnosis of SZ. Furthermore, the association with global negative symptoms suggest a more general role of structural OFC abnormalities in the manifestation of negative symptoms rather than a specific mechanism related to a distinct negative symptom dimension or domain.

It is important to note that some previous studies investigating negative symptoms and structural abnormalities in at-risk stages and first-episode psychosis have been inconclusive.^40,88–90^. As discussed by Walton and colleagues^37^, one possible explanation could be that the association between negative symptoms and OFC thickness is less prominent in earlier disease stages. Additionally, using the PANSS Negative scores rather than specific assessment such as the SANS or newly developed scales (e.g. BNSS, CAINS^91^) could be another explanation for the failure of previous studies to detect an association between OFC thickness and negative symptoms. A different explanation might be that associations between negative symptoms and reduced OFC thickness are only prevalent in specific subgroups of the schizophrenia-spectrum. This could explain why we did not observe the same pattern in individuals with SPT, showing no association with LOFC thickness and even an inverse trend-level association with greater right MOFC thickness in those with higher negative symptoms. In accordance with the latter argument, group comparison revealed a consistent pattern of greater LOFC and MOFC thickness in individuals with SPT compared to HC as well as FEP and SZ patients. A recent report comparing high schizotypy and low schizotypy found reduced volume and thickness predominantly in the temporal and some frontal regions but sparing the OFC.^92^ Interestingly, greater OFC thickness has also been observed in drug-naïve first-episode patients with SZ compared to HC.^88,90^ Our results contribute to this literature, showing thicker OFC in drug-naïve FEP patients compared to medicated patients (FEP, SZ). The nature of preserved or greater prefrontal cortical thickness and volume in early stages of the schizophrenia-spectrum is the subject of intense debate. One prominent hypothesis proposes that preserved/greater prefrontal cortical thickness reflects a compensatory mechanism protecting individuals with SPT from transition to SZ.^38,88,92,93^ A similar mechanism could explain thicker OFC in FEP patients with less severe symptom burden and less need for antipsychotic treatment. Alternatively, greater cortical thickness in specific brain regions could be the consequence of abnormal cortical development during maturation due to insufficient synaptic pruning and/or deficient myelination.^88,94,95^ Such microstructural deficits would therefore contribute to a higher vulnerability in at-risk stages, with SZ only emerging in the presence of additional environmental, biological or genetic factors.^94,96^ Of note, this model does not contradict findings of reduced gray matter in some brain regions. Distinct biological processes may propagate as diverse, heterochronic and distributed effects, thus contributing to the complex neuroanatomical pattern. In this regard, future large-scale longitudinal studies are needed to clarify the origin of these dynamic cortical changes evolving across non-clinical to clinical manifestation within the schizophrenia-spectrum.

### Limitation and future directions

Capitalizing on data from four different sites we aimed to replicate and extend findings on morphometric correlates of negative symptoms for different subgroups of the schizophrenia-spectrum. However, sample sizes within each study and subgroup were rather small and replication in larger cohorts is needed. Negative symptom measures across studies were heterogenous and only one study was a priori designed to investigate the full schizophrenia-spectrum (Zurich dataset) and employed the same negative symptom measures in all subgroups. In addition, this study was the only one that included criteria to minimize the inclusion of secondary negative symptoms related to other sources such as depression.^97,98^ Potential confounding effects of secondary negative symptoms can be assumed in FEP patients from the Basel dataset, where a depressive episode was not an exclusion criterion. In this regard, it will be critical to systematically account for primary and secondary negative symptoms and harmonize psychopathological measures across non-clinical and clinical populations to enable large-scale transdiagnostic approaches crossing boundaries between non-clinical and clinical stages.

## Conclusion

Employing a schizophrenia-spectrum approach, we identified distinct orbitofrontal and striatal structural correlates of negative symptoms in non-clinical/unmedicated populations and early psychosis. Our findings suggest that negative symptoms associate with a temporal continuum of orbitofrontal-striatal abnormalities that may predate the occurrence of schizophrenia. Combining data from four different sites, two divergent neuroanatomical orbitofrontal-striatal patterns emerged along a disease severity axis. One pattern reflected greater striatal volumes in those with chronic disease and exposure to antipsychotic medication, while a second revealed greater OFC thickness in those with subclinical psychotic traits and unmedicated early disease stages.

## Data Availability

All relevant data are
within the paper and its Supporting Information
files. Additional data queries may be directed to Dr Matthias Kirschner

## Conflict of interests

Stefan Kaiser has received speaker honoraria from Roche, Lundbeck, Janssen and Takeda. He receives royalties for cognitive test and training software from Schuhfried. Philippe Tobler has received grant support from Pfizer. Erich Seifritz has received grant support from H. Lundbeck and has served as a consultant and/or speaker for AstraZeneca, Otsuka, Takeda, Eli Lilly, Janssen, Lundbeck, Novartis, Pfizer, Roche, and Servier. None of these activities are related to the present study. All other authors declare no biomedical financial interests or potential conflicts of interest.

## Acknowledgments

Matthias Kirschner acknowledges support from the National Bank Fellowship (McGill University) and the Swiss National Foundation (P2SKP3_178175). The authors thank Dr. Casey Paquola for her helpful insight on the manuscript and excellent support, and all patients and healthy volunteers for their participation.

